# CLINICAL CHARACTERISTICS OF USERS OF WEIGHT LOSS DRUGS: POPULATION-BASED CASE-CONTROL STUDY

**DOI:** 10.64898/2026.02.04.26345477

**Authors:** Inger Johanne Bakken, Paz Lopez-Doriga Ruiz, Kari Furu, Hanne L Gulseth, Kari Anne Sveen, Kjersti Nøkleby, Haakon E. Meyer, Lars J. Kjerpeseth, Øystein Karlstad

## Abstract

**Objectives:** To investigate clinical characteristics of users of weight loss drugs in Norway.

**Design:** Nested population-based case control study

**Setting:** Nationwide healthcare registers in Norway with information on all dispensed medications linked to contacts with primary and specialist healthcare services.

**Participants:** All individuals aged 18-74 years with first dispensing of a weight loss drug (WLD) in 2023–2024, classified as initiators of 1) semaglutide (only Wegovy), 2) liraglutide (only Saxenda), 3) tirzepatide, 4) bupropion–naltrexone, or 5) orlistat. We matched each WLD user by sex and age to five controls randomly selected from the general Norwegian population.

**Main outcome measures:** Type and count of number of comorbidities diagnosed in the two years preceding initiation of WLD use, comedications dispensed in the preceding year, and any previous bariatric surgery. Differences between WLD users and population controls were estimated using conditional logistic regression adjusted for country of birth and education level.

**Results:** During 2023-24, 150,036 individuals initiated semaglutide, 4,603 liraglutide, 3,596 tirzepatide, 31,172 bupropion–naltrexone, and 1,411 orlistat. Among WLD users, 22-29% were registered without any of the pre-defined comorbidities, compared to around half of population controls. Comorbidities and comedications were generally observed at similar proportions in the different WLD user groups, but at much lower proportion for controls: hypertension: 32-38% of WLD users vs. 17-20% of controls; hyperlipidaemias: 20-24% vs. 12-15%; sleep apnoea: 7-11% vs. 2-3%; back pain: 15-20% vs. 10-11%; opioids: 24-30%vs. 13-15%; antidepressants: 20-24%vs. 10-11%.

**Conclusions:** Like other countries, Norway faces the challenge of costly pharmacological obesity treatment, and the need for measures to mitigate increasing socioeconomic health disparities to ensure access to obesity treatment for those most likely to benefit including persons with multimorbidity.

**What is already known on this topic:**

- Obesity is increasing worldwide and is associated with increased risk of multimorbidity.
- The available treatment options for obesity have been rapidly and substantially transformed by the availability of new weight loss drugs.

**What this study adds:**

- This nested case-control study within the Norwegian population examined clinical characteristics of new users of five weight loss drugs: semaglutide, liraglutide, tirzepatide, bupropion–naltrexone, and orlistat. The study identified a higher proportion of weight loss drugs users with cardiovascular, endocrine, mental, and other health conditions, as well as increased usage of cardiovascular, nervous system, and chronic pain medications, compared to the general population.
- The majority of WLD users had high levels of multimorbidity and medication use compared to controls. Approximately 1 in 5 WLD users had no registered comorbidity in either primary or specialist health care systems.
- Additionally, we noted differences in sociodemographic factors between users and population controls, such as lower education levels among WLD users.
- The rapidly expanding use of weight loss drugs necessitates close attention from public health authorities.

## Introduction

Obesity is a growing global health concern and a risk factor for numerous adverse health outcomes.^1^ Individuals with obesity are five times more likely to experience multimorbidity compared to individuals without obesity,^2^ and are at increased risk for premature mortality.^3^ ^4^ Moreover, obesity shares risk factors with other conditions that reduce quality of life,^2^ for instance lack of sleep, chronic stress, poor nutrition, and a sedentary lifestyle.

Obesity contributes to development of chronic diseases including diabetes and endocrine disorders,^5^ cancer,^6^ cardiovascular diseases,^7^ mental health disorders,^8^ musculoskeletal conditions,^9^ and obstructive sleep apnoea.^10^ In a study from the USA, more than 70% of individuals with obesity were affected by multimorbidity.^11^ Obesity is more common in disadvantaged groups, and individuals with lower education and income also experience higher rates of other chronic diseases.^12^ These additional health issues underscore the importance of implementing effective weight loss interventions.^13^ ^14^

New classes of weight loss drugs (WLDs) have changed the treatment of obesity. Incretin-based WLDs (GLP-1 receptor agonists) have demonstrated average weight loss exceeding 12% of initial body weight.^15^ WLDs are increasingly being used as part of obesity management.^16^ However, access to WLDs is restricted by costs and reimbursement practices.^17^ Since 2017, the use of WLDs in Norway has increased markedly, particularly among women.^18^ ^19^

In this real-world study, we aimed to investigate comorbidities and comedication among initiators of WLDs during 2023-2024 within the entire adult Norwegian population. We linked nationwide register data on pharmacy-dispensed drugs with data on diagnoses and surgical procedures from primary and specialist healthcare services.

## Methods

### Available weight loss drugs in Norway

There are currently (2025) five drugs approved and marketed for weight loss: semaglutide (Wegovy), liraglutide (Saxenda), tirzepatide (Mounjaro), bupropion–naltrexone (Mysimba) and orlistat (Xenical). In Norway, reimbursement for weight-loss drugs is highly restrictive and largely limited to bupropion–naltrexone. Monthly WLD supply costs vary from about 75€ (orlistat) and up to about 400€ (tirzepatide). Additional details on market introduction, reimbursement criteria, and pricing in Norway can be found in the Supplemental Material.

### Study population

#### WLD users

The definition of WLD users was based on prescription fills. As semaglutide and liraglutide are also indicated for treatment of type 2 diabetes, we included only products with an approved indication for weight loss. Accordingly, Saxenda and Wegovy were included, but not Victoza, Ozempic and Rybelsus. Tirzepatide is indicated for both weight loss and type 2 diabetes. However, due to its high cost and lack of reimbursement for both conditions, we assumed that tirzepatide predominantly was used for weight loss. For further details refer to Table S1.

Individuals aged 18–74 years who initiated WLDs between January 1^st^ 2023 and December 31^st^ 2024 were included as WLD users. We defined five study populations, each pertaining to initiation of a specific WLD. We used all available data from 1^st^ January 2004 as washout period for prior use of the specific WLD. The index date was the date of the first prescription fill for the specific WLD. I.e. under these criteria, persons included as new users of tirzepatide may have used other types of WLDs at any time prior to initiating tirzepatide.

#### Population controls

For each WLD user, we randomly matched on sex and age to five controls from the general Norwegian population who were alive and residing in Norway on the WLD user’s index date, and who had not previously used that specific WLD. The resulting five analysis data sets consisted of N WLD users and 5*N controls. Within each group consisting of one WLD user and five controls, we assigned the user’s study ID as the group ID and the user’s index date as the group index date. Controls were selected independently for each WLD, meaning that, for instance, a user of tirzepatide who never initiated orlistat could be included among controls for the orlistat study population.

#### Comorbidities and comedications

We established a list of comorbidities and comedications based on previous studies and clinical knowledge,^2^ ^20^ see Supplemental Table 1 for full list with corresponding diagnosis, procedure and drug ATC codes. For comorbidities, we included registrations in the 730 days period prior to the index date. For definition of comedications, the time frame was 365 days (Supplemental Figure 1). For bariatric surgery, we utilized all available data from January 1^st^ 2008 until index date.

We additionally created broader groups of 10 conditions/diseases and counted the number of different conditions registered to define comorbidity level (categorized as none, 1-3 and 4 comorbidities). For details, see Supplemental Table 1.

#### Data sources

We retrieved information on year of birth, sex and residential status (dates of birth, death, emigration and immigration) and education on the highest achieved level in 2023 from Statistics Norway. Education was categorized as ≤ 10 years (primary), 11-13 years (secondary) and ≥14 years (tertiary). Country of birth was categorized as Norway, Europe outside Norway, and outside Europe.

Data on prescription fills were retrieved from the Norwegian Prescribed Drug Registry (NorPD). This registry holds individual-level information on all prescription fills at Norwegian pharmacies from 2004 onwards.^21^

Data on diagnoses (ICD-10) and surgical procedures in specialist health care were from the Norwegian Patient Registry (NPR), while data on diagnoses (ICPC2) in primary health care were from the Norwegian Registry for Primary Health Care (NRPHC).^22^

The encrypted personal identity number unique to every Norwegian citizen was used to link data across data sources.

### Statistical analysis

We used logistic regression to summarise associations between comorbidity/comedication and WLD use, adjusting for age, sex, education and country of birth.^23^ All data handling and analysis was carried out using the statistical software R, version 4.3.0.^24^

## Results

From 2016 to 2024, there was a large increase in the number of individuals using WLDs, accompanied by a large shift in the distribution of medications used (Figure 1). In 2024, the total number of WLD users was nearly 164,000, corresponding to 4.1% of the Norwegian population 18-74 years old. The highest proportion was seen among women 50-59 years old (9.6%). During 2023–2024, semaglutide was the most common WLD (Figure 1, Table 1).

**Figure 1.**
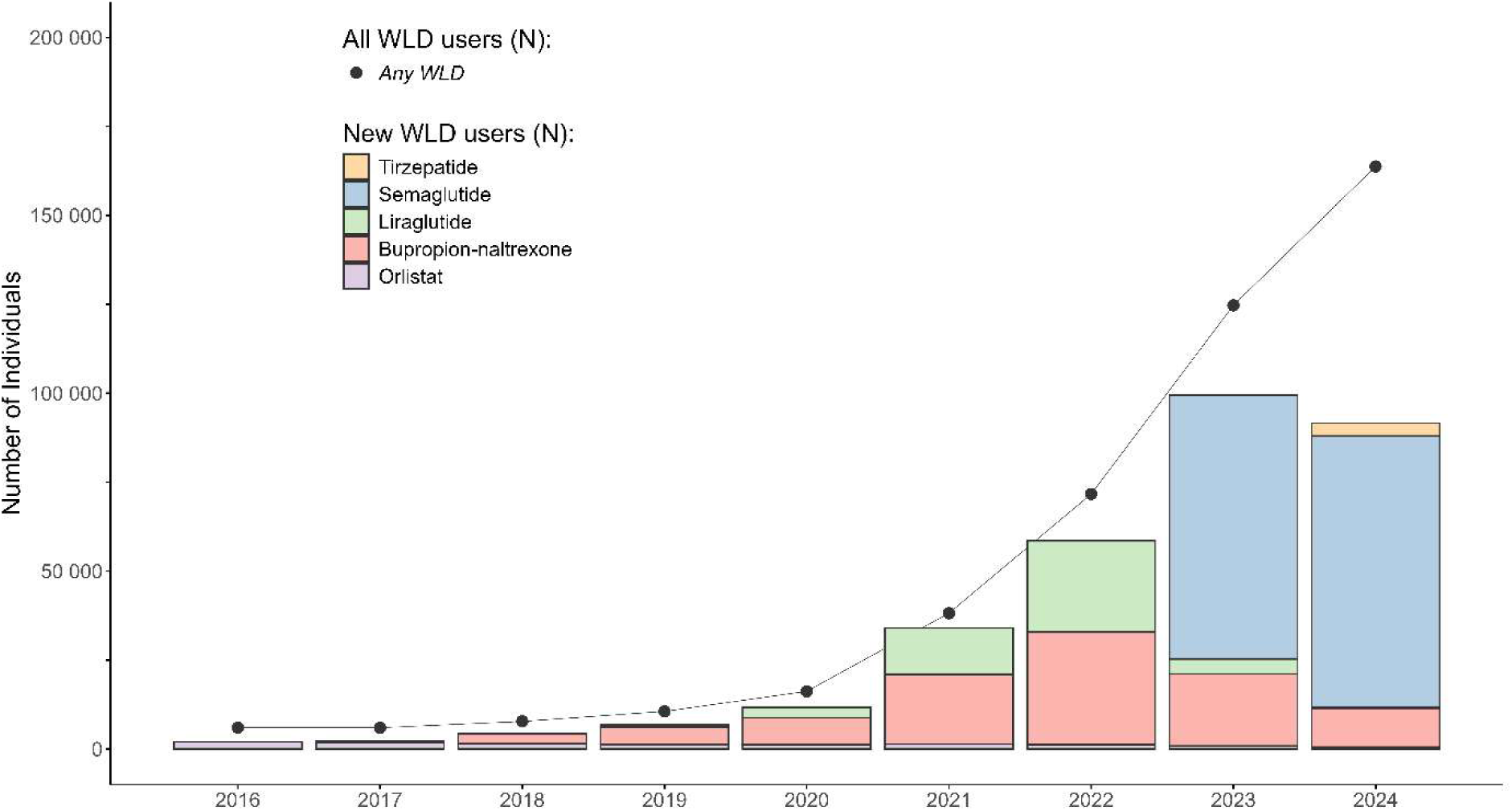
Norway 2016-2024. Annual number of weight loss drugs (grey line). New users by type of drug as coloured boxes. New users in 2023-2024 were included in further analyses.

**Table 1.**
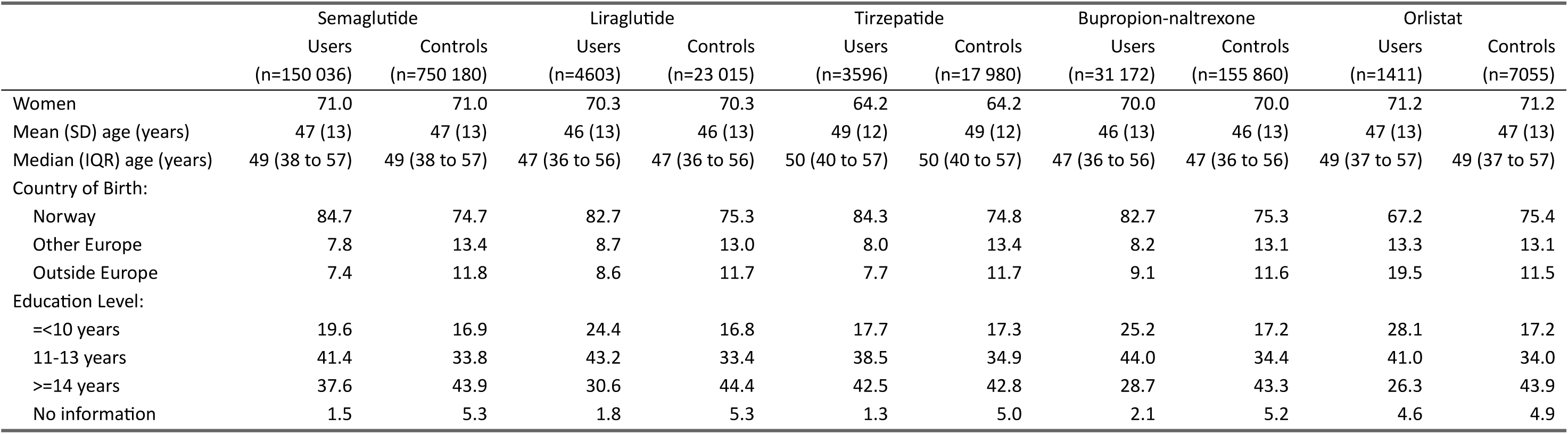
New users of weight loss drugs compared to controls, Norway 2023-2024. Controls matched by sex and age. Percentages unless otherwise stated.

|  | Semaglutide |  | Liraglutide |  | Tirzepatide |  | Bupropion-naltrexone |  | Orlistat |  |
| --- | --- | --- | --- | --- | --- | --- | --- | --- | --- | --- |
|  | Users | Controls | Users | Controls | Users | Controls | Users | Controls | Users | Controls |
|  | (n=150 036) | (n=750 180) | (n=4603) | (n=23 015) | (n=3596) | (n=17 980) | (n=31 172) | (n=155 860) | (n=1411) | (n=7055) |
| Women | 71.0 | 71.0 | 70.3 | 70.3 | 64.2 | 64.2 | 70.0 | 70.0 | 71.2 | 71.2 |
| Mean (SD) age (years) | 47 (13) | 47 (13) | 46 (13) | 46 (13) | 49 (12) | 49 (12) | 46 (13) | 46 (13) | 47 (13) | 47 (13) |
| Median (IQR) age (years) | 49 (38 to 57) | 49 (38 to 57) | 47 (36 to 56) | 47 (36 to 56) | 50 (40 to 57) | 50 (40 to 57) | 47 (36 to 56) | 47 (36 to 56) | 49 (37 to 57) | 49 (37 to 57) |
| Country of Birth: |  |  |  |  |  |  |  |  |  |  |
| Norway | 84.7 | 74.7 | 82.7 | 75.3 | 84.3 | 74.8 | 82.7 | 75.3 | 67.2 | 75.4 |
| Other Europe | 7.8 | 13.4 | 8.7 | 13.0 | 8.0 | 13.4 | 8.2 | 13.1 | 13.3 | 13.1 |
| Outside Europe | 7.4 | 11.8 | 8.6 | 11.7 | 7.7 | 11.7 | 9.1 | 11.6 | 19.5 | 11.5 |
| Education Level: |  |  |  |  |  |  |  |  |  |  |
| <=10 years | 19.6 | 16.9 | 24.4 | 16.8 | 17.7 | 17.3 | 25.2 | 17.2 | 28.1 | 17.2 |
| 11-13 years | 41.4 | 33.8 | 43.2 | 33.4 | 38.5 | 34.9 | 44.0 | 34.4 | 41.0 | 34.0 |
| >=14 years | 37.6 | 43.9 | 30.6 | 44.4 | 42.5 | 42.8 | 28.7 | 43.3 | 26.3 | 43.9 |
| No information | 1.5 | 5.3 | 1.8 | 5.3 | 1.3 | 5.0 | 2.1 | 5.2 | 4.6 | 4.9 |

While more women than men used WLDs overall, the proportion of women using tirzepatide was lower at 64%, compared 70-71% for other WLDs. The median age at first prescription fill varied slightly between the WLDs (47–50 years). For WLDs other than orlistat, a higher proportion of users than population controls were born in Norway. For orlistat, the proportion born in Norway was substantially lower than for other WLDs and was also below the proportion among controls. For WLDs other than tirzepatide, the proportion having higher education was lower among users compared to their respective controls. For tirzepatide, we observed no differences between users and controls regarding education levels.

### Overweight and obesity, prior bariatric surgery and prior WLD use

Among WLD users, 55–77% had a registered overweight or obesity diagnosis, in contrast to 5–10% of controls (Table 2). We observed a higher frequency of bariatric surgery among WLD users compared to controls.

**Table 2.** Overweight and obesity diagnosis, prior bariatric surgery, prior use of other weight loss drugs, and number of comorbidities among users of weight loss drugs and controls. Controls matched by sex and age. Percentages unless otherwise stated. Norway 2023-2024.

|  | Semaglutide |  | Liraglutide |  | Tirzepatide |  | Bupropion-naltrexone |  | Orlistat |  |
| --- | --- | --- | --- | --- | --- | --- | --- | --- | --- | --- |
|  | Users | Controls | Users | Controls | Users | Controls | Users | Controls | Users | Controls |
|  | (n=150 036) | (n=750 180) | (n=4603) | (n=23 015) | (n=3596) | (n=17 980) | (n=31 172) | (n=155 860) | (n=1411) | (n=7055) |
| Overweight/obesity diagnosis | 63 | 4.6 | 69 | 5.9 | 77 | 9.5 | 61 | 6.5 | 55 | 7.1 |
| Prior bariatric surgery | 3.3 | 0.8 | 4.1 | 0.9 | 3.6 | 1.0 | 3.3 | 0.9 | 1.5 | 1.0 |
| Other WLDs in prior year |  |  |  |  |  |  |  |  |  |  |
| Any AOM | 20 | 1.5 | 37 | 1.9 | 74 | 6.0 | 9.1 | 2.8 | 27 | 4.4 |
| Semaglutide | - | - | 3.7 | 0.6 | 70 | 4.8 | 4.9 | 1.8 | 6.1 | 2.3 |
| Liraglutide | 9.6 | 0.6 | - | - | 3.9 | 0.4 | 3.8 | 1.1 | 7.5 | 1.2 |
| Bupropion-naltrexone | 12 | 0.9 | 33 | 1.3 | 6.4 | 1.0 | - | - | 16 | 1.3 |
| Orlistat | 0.7 | 0.1 | 1.9 | 0.1 | 0.6 | 0.1 | 0.8 | 0.1 | - | - |
| Comorbidity level |  |  |  |  |  |  |  |  |  |  |
| None | 29 | 51 | 24 | 52 | 26 | 49 | 27 | 51 | 22 | 51 |
| 1-3 | 52 | 40 | 53 | 39 | 52 | 41 | 53 | 40 | 54 | 40 |
| >= 4 | 19 | 9 | 24 | 8 | 22 | 10 | 20 | 9 | 24 | 9 |
| Number of Comorbidities: |  |  |  |  |  |  |  |  |  |  |
| Median (IQR) | 2 (0-3) | 0 (0-2) | 2 (1-3) | 0 (0-2) | 2 (0-3) | 1 (0-2) | 2 (0-3) | 0 (0-2) | 2 (1-3) | 0 (0-2) |

A substantial proportion of individuals initiating a specific WLD had used other types of WLDs in the year preceding the index date. This was especially common among new users of tirzepatide, with nearly three-quarters having tried other WLD in the preceding year.

### Comorbidities and comedications

A high comorbidity level (four or more comorbidities) was more than twice as common in WLD users (19–24%) as in controls (8–10%) (Table 2). Whereas 22–24% of orlistat and liraglutide users and 26–28% of users of the other WLDs had none of the predefined comorbidities, this was the case for half of controls.

The use of WLDs was strongly associated with most of the individual comorbidities studied (Table 3), with similar patterns for all WLDs. Hypertension had the highest proportion across all WLDs and was observed approximately twice as often in WLD users as in controls. For all WLDs, the strongest odds ratio for difference in comorbidity among users and controls was for sleep apnoea. Hyperlipidaemia was also more common in WLD users than in controls, as was back pain and arthrosis.

**Table 3.** Proportion (%) and Odds Ratio (OR) of registered comorbidities among users of weight loss drugs and population controls. Norway 2023-2024.

|  | Semaglutide |  |  | Liraglutide |  |  | Tirzepatide |  |  | Bupropion-naltrexone |  |  | Orlistat |  |  |
| --- | --- | --- | --- | --- | --- | --- | --- | --- | --- | --- | --- | --- | --- | --- | --- |
|  | User (%) | Control (%) | OR (95% CI) | User (%) | Control (%) | OR (95% CI) | User (%) | Control (%) | OR (95% CI) | User (%) | Control (%) | OR (95% CI) | User (%) | Control (%) | OR (95% CI) |
| Hypertension | 32 | 17 | 2.5<br>(2.5 to 2.5) | 35 | 17 | 3.0<br>(2.8 to 3.3) | 38 | 20 | 2.8<br>(2.5 to 3.0) | 33 | 17 | 2.6<br>(2.5 to 2.7) | 37 | 19 | 2.8<br>(2.4 to 3.2) |
| Heart failure | 0.8 | 0.5 | 1.6<br>(1.5 to 1.7) | 1.2 | 0.4 | 2.6<br>(1.8 to 3.6) | 1.0 | 0.5 | 2.1<br>(1.4 to 3.1) | 0.7 | 0.5 | 1.3<br>(1.1 to 1.5) | 1.0 | 0.6 | 1.6<br>(0.8 to 3.1) |
| Ischaemic heart disease | 2.6 | 1.6 | 1.6<br>(1.6 to 1.7) | 3.5 | 1.6 | 2.2<br>(1.8 to 2.7) | 3.6 | 1.9 | 2.1<br>(1.7 to 2.6) | 2.7 | 1.6 | 1.6<br>(1.5 to 1.7) | 3.7 | 1.6 | 2.1<br>(1.4 to 2.9) |
| Atrial fibrillation | 1.7 | 0.9 | 2.0<br>(1.9 to 2.1) | 2.1 | 0.9 | 2.5<br>(1.9 to 3.2) | 2.0 | 0.9 | 2.2<br>(1.7 to 3.0) | 1.4 | 0.9 | 1.5<br>(1.4 to 1.7) | 1.6 | 0.8 | 2.0<br>(1.2 to 3.4) |
| T1D | 0.6 | 0.9 | 0.6<br>(0.6 to 0.7) | 1.7 | 1.0 | 1.6<br>(1.2 to 2.1) | 0.7 | 0.9 | 0.8<br>(0.5 to 1.2) | 1.4 | 0.9 | 1.4<br>(1.3 to 1.6) | 1.6 | 1.0 | 1.6<br>(1.0 to 2.5) |
| T2D | 3.8 | 4.3 | 0.9<br>(0.9 to 0.9) | 5.5 | 4.1 | 1.3<br>(1.2 to 1.6) | 6.5 | 4.9 | 1.4<br>(1.2 to 1.7) | 6.2 | 4.1 | 1.4<br>(1.4 to 1.5) | 7.6 | 4.1 | 1.7<br>(1.3 to 2.1) |
| Hyper-lipidaemias | 20 | 12 | 1.8<br>(1.8 to 1.9) | 21 | 12 | 2.1<br>(2.0 to 2.4) | 24 | 15 | 1.9<br>(1.7 to 2.1) | 20 | 12 | 1.9<br>(1.8 to 2.0) | 23 | 13 | 2.1<br>(1.8 to 2.5) |
| ADHD | 3.5 | 2.2 | 1.4<br>(1.4 to 1.4) | 4.5 | 2.3 | 1.7<br>(1.4 to 2.0) | 3.9 | 2.4 | 1.4<br>(1.2 to 1.8) | 3.9 | 2.4 | 1.3<br>(1.3 to 1.4) | 4.3 | 2.3 | 1.7<br>(1.2 to 2.3) |
| Anxiety | 6.3 | 4.2 | 1.4<br>(1.4 to 1.4) | 8.7 | 4.6 | 1.7<br>(1.5 to 2.0) | 7.6 | 4.6 | 1.6<br>(1.4 to 1.9) | 6.7 | 4.5 | 1.3<br>(1.3 to 1.4) | 8.3 | 4.4 | 1.9<br>(1.5 to 2.4) |
| Depression | 13 | 7.3 | 1.7<br>(1.7 to 1.7) | 19 | 8.2 | 2.4<br>(2.2 to 2.6) | 17 | 8.0 | 2.3<br>(2.1 to 2.5) | 14 | 8.1 | 1.7<br>(1.7 to 1.8) | 16 | 7.7 | 2.0<br>(1.7 to 2.4) |
| Drug-related disorders | 2.2 | 1.9 | 1.0<br>(0.9 to 1.0) | 3.5 | 2.0 | 1.5<br>(1.2 to 1.7) | 2.8 | 2.1 | 1.2<br>(1.0 to 1.5) | 2.1 | 2.0 | 0.8<br>(0.8 to 0.9) | 4.1 | 1.8 | 2.0<br>(1.4 to 2.7) |
| Sleep apnoea | 7.0 | 1.9 | 3.9<br>(3.8 to 4.0) | 11 | 2.0 | 5.7<br>(5.0 to 6.5) | 11 | 2.6 | 4.5<br>(3.9 to 5.2) | 8.7 | 2.1 | 4.4<br>(4.1 to 4.6) | 7.8 | 2.3 | 3.3<br>(2.5 to 4.3) |
| Stress incontinence | 3.1 | 1.7 | 1.8<br>(1.7 to 1.9) | 3.6 | 1.8 | 1.9<br>(1.6 to 2.3) | 4.1 | 2.3 | 1.8<br>(1.5 to 2.2) | 3.6 | 1.8 | 1.9<br>(1.8 to 2.1) | 4.3 | 1.9 | 2.2<br>(1.6 to 3.0) |
| Arthrosis | 9.6 | 4.7 | 2.2<br>(2.2 to 2.2) | 12 | 5.3 | 2.4<br>(2.2 to 2.7) | 11 | 5.9 | 2.1<br>(1.8 to 2.4) | 11 | 5.0 | 2.4<br>(2.3 to 2.5) | 11 | 5.7 | 1.8<br>(1.5 to 2.3) |
| Back pain | 15 | 10 | 1.7<br>(1.6 to 1.7) | 19 | 10 | 1.9<br>(1.8 to 2.1) | 20 | 11 | 2.0<br>(1.8 to 2.2) | 18 | 10 | 1.8<br>(1.8 to 1.9) | 20 | 10 | 1.9<br>(1.6 to 2.2) |

Odds ratio (OR) with 95% Confidence intervals (CI) from logistic regression, adjusted for age, sex, country of birth and level of education.

The high degree of somatic comorbidities among WLD users was also reflected in the comedication patterns (Table 4). For instance, among initiators of bupropion–naltrexone, approximately one third had used antihypertensive medications compared to one sixth of controls. Among tirzepatide initiators, half had used anti-inflammatory medications (mainly NSAIDs), while this was the case for one in four of controls. Further, use of opioids was substantially higher among all WLD users than controls, e.g. 35% of tirzepatide initiators had used opioids during the last year compared to 15% of their respective controls.

**Table 4.**
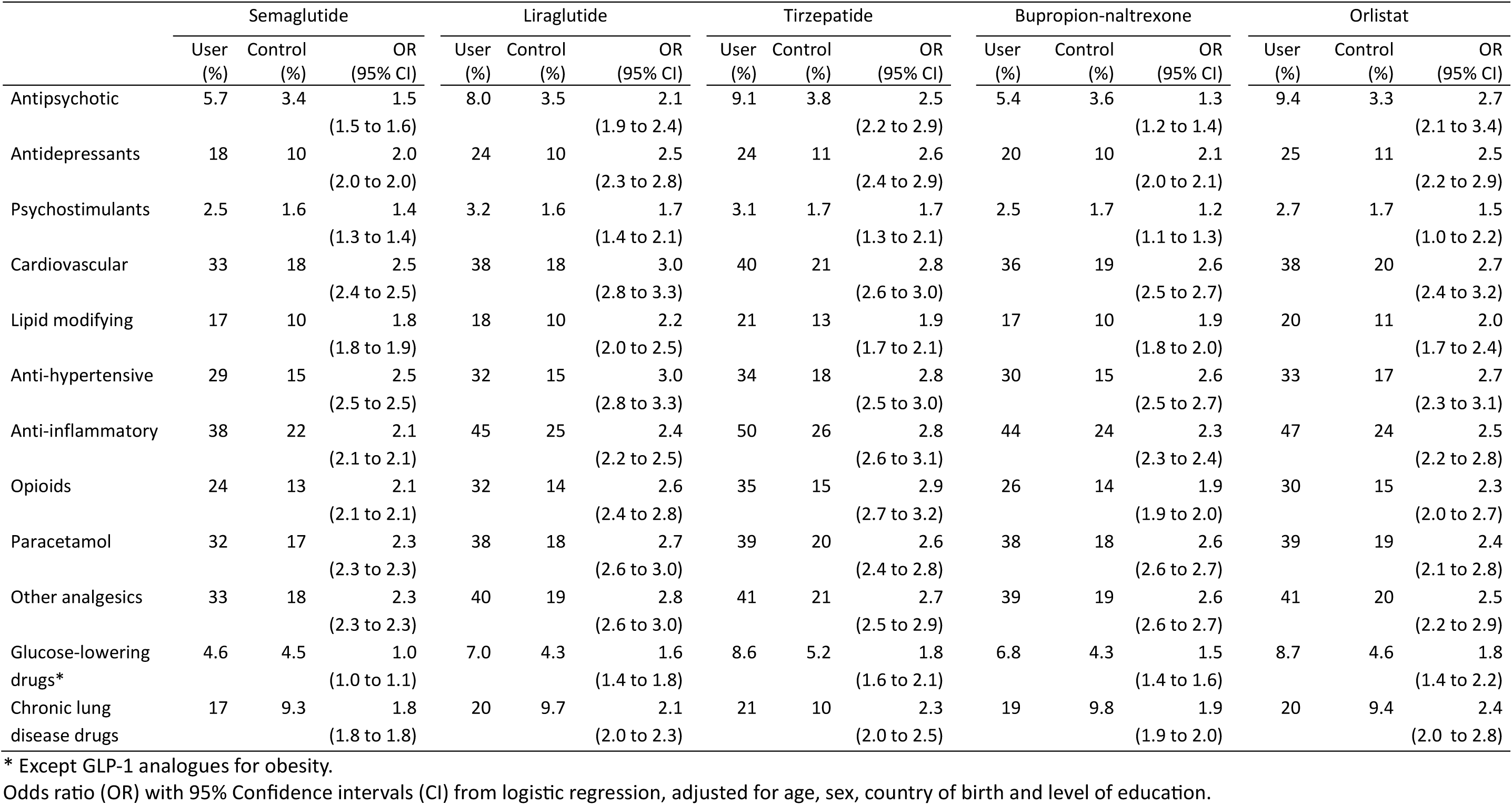
Proportion (%) and Odds Ratio (OR) of registered comedications among users of weight loss drugs and population controls. Norway 2023-2024.

|  | Semaglutide |  |  | Liraglutide |  |  | Tirzepatide |  |  | Bupropion-naltrexone |  |  | Orlistat |  |  |
| --- | --- | --- | --- | --- | --- | --- | --- | --- | --- | --- | --- | --- | --- | --- | --- |
|  | User (%) | Control (%) | OR (95% CI) | User (%) | Control (%) | OR (95% CI) | User (%) | Control (%) | OR (95% CI) | User (%) | Control (%) | OR (95% CI) | User (%) | Control (%) | OR (95% CI) |
| Antipsychotic | 5.7 | 3.4 | 1.5<br>(1.5 to 1.6) | 8.0 | 3.5 | 2.1<br>(1.9 to 2.4) | 9.1 | 3.8 | 2.5<br>(2.2 to 2.9) | 5.4 | 3.6 | 1.3<br>(1.2 to 1.4) | 9.4 | 3.3 | 2.7<br>(2.1 to 3.4) |
| Antidepressants | 18 | 10 | 2.0<br>(2.0 to 2.0) | 24 | 10 | 2.5<br>(2.3 to 2.8) | 24 | 11 | 2.6<br>(2.4 to 2.9) | 20 | 10 | 2.1<br>(2.0 to 2.1) | 25 | 11 | 2.5<br>(2.2 to 2.9) |
| Psychostimulants | 2.5 | 1.6 | 1.4<br>(1.3 to 1.4) | 3.2 | 1.6 | 1.7<br>(1.4 to 2.1) | 3.1 | 1.7 | 1.7<br>(1.3 to 2.1) | 2.5 | 1.7 | 1.2<br>(1.1 to 1.3) | 2.7 | 1.7 | 1.5<br>(1.0 to 2.2) |
| Cardiovascular | 33 | 18 | 2.5<br>(2.4 to 2.5) | 38 | 18 | 3.0<br>(2.8 to 3.3) | 40 | 21 | 2.8<br>(2.6 to 3.0) | 36 | 19 | 2.6<br>(2.5 to 2.7) | 38 | 20 | 2.7<br>(2.4 to 3.2) |
| Lipid modifying | 17 | 10 | 1.8<br>(1.8 to 1.9) | 18 | 10 | 2.2<br>(2.0 to 2.5) | 21 | 13 | 1.9<br>(1.7 to 2.1) | 17 | 10 | 1.9<br>(1.8 to 2.0) | 20 | 11 | 2.0<br>(1.7 to 2.4) |
| Anti-hypertensive | 29 | 15 | 2.5<br>(2.5 to 2.5) | 32 | 15 | 3.0<br>(2.8 to 3.3) | 34 | 18 | 2.8<br>(2.5 to 3.0) | 30 | 15 | 2.6<br>(2.5 to 2.7) | 33 | 17 | 2.7<br>(2.3 to 3.1) |
| Anti-inflammatory | 38 | 22 | 2.1<br>(2.1 to 2.1) | 45 | 25 | 2.4<br>(2.2 to 2.5) | 50 | 26 | 2.8<br>(2.6 to 3.1) | 44 | 24 | 2.3<br>(2.3 to 2.4) | 47 | 24 | 2.5<br>(2.2 to 2.8) |
| Opioids | 24 | 13 | 2.1<br>(2.1 to 2.1) | 32 | 14 | 2.6<br>(2.4 to 2.8) | 35 | 15 | 2.9<br>(2.7 to 3.2) | 26 | 14 | 1.9<br>(1.9 to 2.0) | 30 | 15 | 2.3<br>(2.0 to 2.7) |
| Paracetamol | 32 | 17 | 2.3<br>(2.3 to 2.3) | 38 | 18 | 2.7<br>(2.6 to 3.0) | 39 | 20 | 2.6<br>(2.4 to 2.8) | 38 | 18 | 2.6<br>(2.6 to 2.7) | 39 | 19 | 2.4<br>(2.1 to 2.8) |
| Other analgesics | 33 | 18 | 2.3<br>(2.3 to 2.3) | 40 | 19 | 2.8<br>(2.6 to 3.0) | 41 | 21 | 2.7<br>(2.5 to 2.9) | 39 | 19 | 2.6<br>(2.6 to 2.7) | 41 | 20 | 2.5<br>(2.2 to 2.9) |
| Glucose-lowering drugs* | 4.6 | 4.5 | 1.0<br>(1.0 to 1.1) | 7.0 | 4.3 | 1.6<br>(1.4 to 1.8) | 8.6 | 5.2 | 1.8<br>(1.6 to 2.1) | 6.8 | 4.3 | 1.5<br>(1.4 to 1.6) | 8.7 | 4.6 | 1.8<br>(1.4 to 2.2) |
| Chronic lung disease drugs | 17 | 9.3 | 1.8<br>(1.8 to 1.8) | 20 | 9.7 | 2.1<br>(2.0 to 2.3) | 21 | 10 | 2.3<br>(2.0 to 2.5) | 19 | 9.8 | 1.9<br>(1.9 to 2.0) | 20 | 9.4 | 2.4<br>(2.0 to 2.8) |

\* Except GLP-1 analogues for obesity.
Odds ratio (OR) with 95% Confidence intervals (CI) from logistic regression, adjusted for age, sex, country of birth and level of education.

Across all WLDs studied, the proportions of WLD users with a depression diagnosis was about twice as high as that of their controls (Table 3). The pattern of increased levels of depression among WLD users was also evident from antidepressants use (Table 4). WLD users also showed increased levels of anxiety and ADHD, though patterns for drug-related disorders were less clear. Additionally, we observed more use of antipsychotic drugs among WLD users than in control groups.

These patterns were largely consistent across categories of sex and age (Supplemental Table 2).

## Discussion

Using nationwide register data in a nested case-control design with sex and age matching, we compared clinical characteristics of new WLD users to general population controls. The comprehensive coverage of primary care and specialist care healthcare system encounters, along with information on all pharmacy-dispensed drugs, enabled us to study comorbidity and comedication patterns. More women than men used WLDs and the median age at initiation ranged from 47 to 50 years. There were differences in education level and country of birth between users of older versus newer and more expensive WLDs, and between WLD users and controls. Up to a quarter of WLD users were registered with a high comorbidity level, compared to one in ten of controls. Overall, we observed increased frequencies of somatic and mental comorbidities, and higher levels of use of other medications among WLD users compared to controls.

### Sociodemographic characteristics

Obesity is more common in disadvantaged groups, and individuals with lower education and income also experience higher rates of other chronic diseases.^12^ Studies from Scandinavia and the USA have shown higher obesity prevalences in groups with lower socioeconomic status,^12^ ^25^ ^26^ but also less use of WLDs in such groups.^27^ ^28^ A recent review showed that lower income is associated with reduced treatment with GLP-1 receptor agonists.^29^ In our study, a lower proportion of WLD users than controls were foreign-born, except for users of orlistat. Also, users of reimbursable WLDs (orlistat, bupropion–naltrexone and liraglutide) were more likely to have only primary-level education than users of the non-reimbursed, more effective, and more expensive WLDs, indicating a socioeconomic health gap in choice of medication.

### Overweight/obesity and prior WLD use

Prevalence estimates of overweight and obesity cannot be based on registry data, as diagnoses for overweight or obesity were registered for 5 to 10% of controls only, far below survey-based estimates.^30^ Among WLD users, the proportion registered with overweight and obesity diagnoses during the two years before WLD initiation ranged from 55% for orlistat to 77% for tirzepatide.

A substantial proportion of individuals initiating a WLD had tried other WLDs in the previous year, probably reflecting side effects and switches to new and more effective drugs when these became available on the market in 2023 (semaglutide/Wegovy) and 2024 (tirzepatide). A recent study from the USA showed high rates of discontinuation and reinitiations, largely due to side effects and costs.^31^

### Comorbidity patterns

Obesity is associated with numerous comorbid conditions and obesity-related complications. A high comorbidity level was more than twice as common in WLD users (19–24%) compared to controls (8–10%), supported by substantially higher levels of most individual conditions and medications studied.

A study from the USA among users of GLP-1 receptor agonists with and without diabetes showed higher proportions of hypertension and hyperlipidaemia, as well as atrial fibrillation, compared to our study. They reported similar rates of depression and ischaemic heart disease.^31^

We observed high levels of obstructive sleep apnoea among WLD users; obesity is a well-known risk factor. A weight gain of 10% has been shown to be associated with a 32% increase in sleep apnoea severity, while a 10% weight loss was associated with 26% reduction in severity.^32^ In the USA, tirzepatide is indicated for sleep apnoea, and individuals who achieve a weight reduction of 18 to 20% may experience remission of the condition.^33^ We observed that the proportions using medications to treat chronic pain were much higher among users of WLD compared to controls, which is line with the three-fold higher prevalence of pain among people with obesity described previously.^34^

### Implications

The majority of WLD users in our study had high levels of multimorbidity and medication use compared to controls, implying that users of WLDs experience considerable somatic and mental health challenges. Severe obesity is a public health problem and should be treated as a chronic condition.^35^ The WHO has recently released the first guidelines on the use of GLP-1 receptor agonists for treatment of obesity.^36^ Weight loss may be an important strategy to prevent or reduce comorbidity and complications among individuals with obesity.^2^ Obesity is more common in disadvantaged groups, and individuals with lower education and income also experience higher rates of other chronic diseases. More options for obesity treatment are now available, but the high cost of the new effective WLDs is still a barrier. In our study, we observed socioeconomic differences between WLD users and controls, and between different WLD groups. As of 2025 only the less effective WLDs bupropion–naltrexone and orlistat are reimbursed for individuals with BMI≥35 with comorbidities in Norway, possibly exacerbating existing socioeconomic disparities. Bupropion–naltrexone is under review by the European Medicine Agency due to cardiovascular risk concerns,^37^ and yearly follow-up of users is recommended.^36^ Policy approaches should be aimed at addressing equal access to obesity treatment for those who are most likely to benefit, irrespective of sociodemographic status.^36^ Obesity is more common in disadvantaged groups, and individuals with lower education and income also experience higher rates of other chronic diseases. Policy approaches should be aimed at addressing equal access to obesity treatment for those who are most likely to benefit, irrespective of sociodemographic status. Obesity is a public health problem and should be treated as a chronic disease.^35^ The WHO has recently released the first guidelines on the use of GLP-1 receptor agonists for treatment of obesity.^36^ Weight loss may be an important strategy to prevent or reduce comorbidity and complications among individuals with obesity.^2^ More options for obesity treatment are now available, but the high cost of the new effective WLDs is still a barrier.^38^ Obesity is more common in disadvantaged groups, and individuals with lower education and income also experience higher rates of other chronic diseases. Policy approaches should be aimed at addressing equal access to obesity treatment for those who are most likely to benefit, irrespective of sociodemographic status. Obesity is a public health problem and should be treated as a chronic disease.^35^ The WHO has recently released the first guidelines on the use of GLP-1 receptor agonists for treatment of obesity.^36^ Weight loss may be an important strategy to prevent or reduce comorbidity and complications among individuals with obesity.^2^ More options for obesity treatment are now available, but the high cost of the new effective WLDs is still a barrier.^38^ Obesity is more common in disadvantaged groups, and individuals with lower education and income also experience higher rates of other chronic diseases. Policy approaches should be aimed at addressing equal access to obesity treatment for those who are most likely to benefit, irrespective of sociodemographic status.^2^ ^35^ ^36^ ^38^ Policy approaches should be aimed at addressing equal access to obesity treatment for those who are most likely to benefit, irrespective of sociodemographic status.

Interestingly, approximately one in five WLD users in our study had no pre-registered comorbidity in primary or specialist health care potentially suggesting use for non-medical weight management rather than for disease prevention.

### Strengths and weaknesses of the study

A major strength of our study is the nationwide access to individual to level data on medication use and the possibility to link comprehensive information on diagnoses and procedures from both primary and specialist healthcare. The mandatory health registers cover all government-financed healthcare in the country thus covering almost all healthcare services use for chronic conditions. The unique person identification number assigned to all residents at birth or immigration ensures precise person-level linkage between registers and over time.

The main limitation was the lack of access to anthropometric measures such as body mass index (BMI), and we are not able to assess weight of WLD users and its association with these comorbidities. Secondly, relying on diagnosis codes from the two years preceding initiation of WLDs can lead to underestimated proportions of comorbidities, as some patients with chronic diseases might not have had follow-up care within these two years. Our data include government-financed healthcare from the primary and secondary healthcare sector. Although most healthcare in Norway is provided through this system, 30 to 40% of bariatric surgeries are performed privately,^39^ and, thus, are not included in our data.

## Conclusion

The use of weight loss drugs is rapidly expanding and a majority of users of these drugs have complex comorbidity patterns. Like other countries with a universal health system, Norway faces the challenge of funding costly pharmacological treatment of obesity and the need for measures to mitigate increasing socioeconomic health disparities, to ensure access to obesity treatment for those most likely to benefit.

## Declarations

### Patient and public involvement

This research project was funded by the Norwegian Institute of Public Health, and no funds were available to involve patients in the research design. However, several of the authors are active clinicians and their interactions with patients inform the conduct and interpretation. Results will be disseminated to the public through media, and to other governmental health institutions through official channels. It is not allowed nor possible for the research group to reidentify and contact study participants identified in the health register data.

### Author Contributions

IJB, PLDR, and ØK designed the study. IJB and PLDR and had the main responsibility for preparation of the manuscript. IJB prepared the data and conducted the analyses. All authors repeatedly provided feedback on the manuscript and approved the final version for submission. KF, HLG, KAS, KN, HEM, LJK, ØK provided information necessary to define comorbidities and comedications. HLG, KAS, KN, HEM, and LK contributed to the clinical interpretation. PLDR, IJB and ØK is the study guarantors. The corresponding author attests that all listed authors meet authorship criteria and that no others meeting the criteria have been omitted.

### Transparency statement

PLDR, IJB and ØK affirm that this manuscript is an honest, accurate, and transparent account of the study being reported; that no important aspects of the study have been omitted; and that any discrepancies from the study as planned have been explained.

### Ethical approval

This research was approved by the Committee for Medical and Health Research Ethics in Norway (REK South to East A; 2017/2546). The requirement to obtain informed consent from research subjects is waived for observational, register-based research in Norway.

### Data Availability Statement

No additional data available. Data use agreements are required from the Norwegian Health Data Access Body (HDS) for accessing health register data, and separately from Statistics Norway to access sociodemographic register data. These agreements do not allow the authors to share the person-level data with researchers who are not covered by the data use agreements. The data is accessible for authorized researchers after receiving ethical approval and applying for data access to helsedata.no and ssb.no/en/data-til-forskning/utlan-av-data-til-forskere.

## Disclaimer

Data from the Norwegian healthcare registers have been used in this publication. The interpretation and reporting of these data are the sole responsibility of the authors, and no endorsement by the healthcare registers is intended or should be inferred.

