## Supplementary Material for "CLINICAL CHARACTERISTICS OF USERS OF WEIGHT LOSS DRUGS: POPULATION-BASED CASE-CONTROL STUDY"

### Supplemental Material

### Figures

#### Figure S1. Study design

For comorbidities we included events during the period from 730 days prior to the index date up to the index date, while we used the previous 365 days for definition of comedications. For bariatric surgery, we used the entire available data period on surgical codes prior to the index date (i.e. from January 1, 2008).

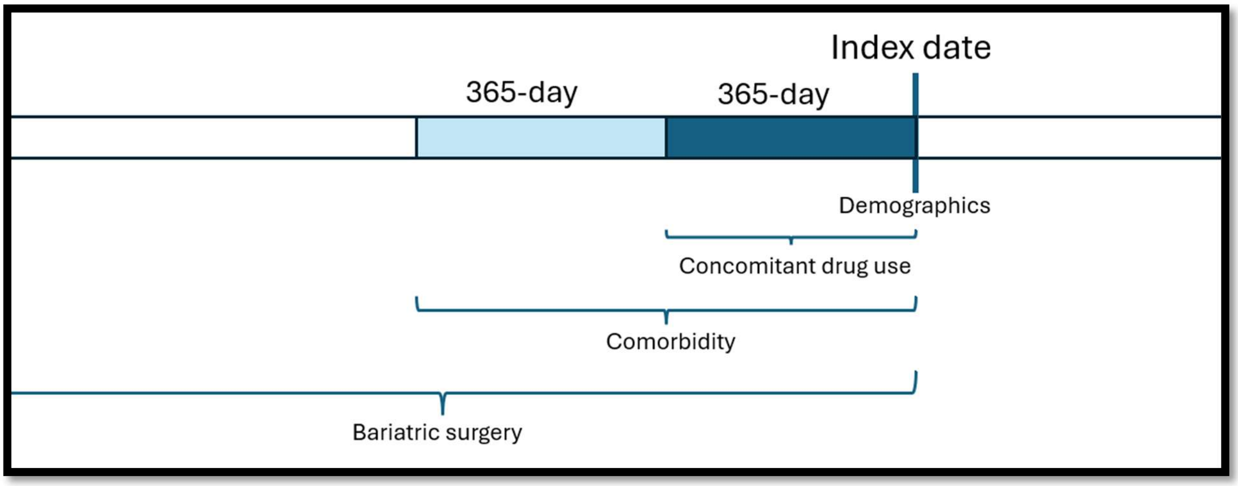

#### Figure S2. Timeline for introduction of weight loss drugs in Norway

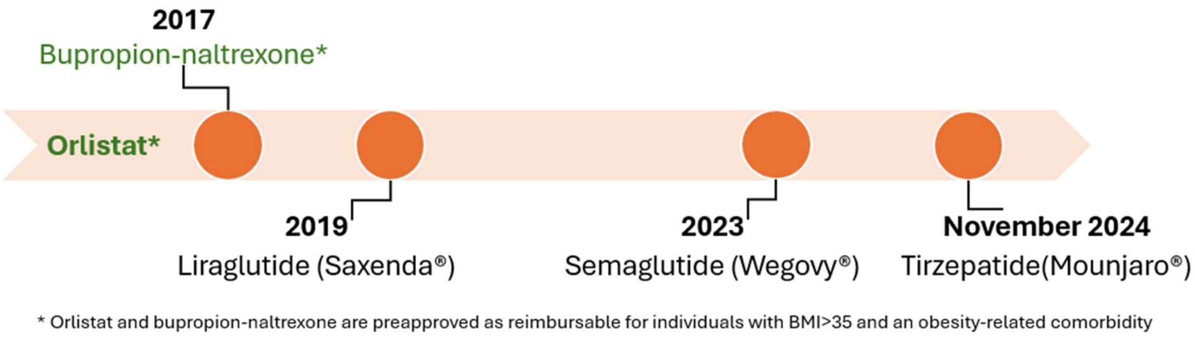

### Tables

#### Table S1. Definitions for weight loss drugs, comorbidity groups, comedication groups and comorbidity counts

| | |
|---|---|
| **WEIGHT LOSS DRUGS** | |
| Semaglutide for diabetes treatment (Ozempic) | ATC: A10BJ06 **AND** Item number*: 178275, 203650, 219433, 226671, 233428, 234460, 259635, 269415, 275621, 279566, 302179, 306901, 315075, 320518, 327000, 345700, 354711, 360141, 363529, 365534, 366288, 406340, 413340, 585776 |
| Orlistat (Xenical) | ATC: A08AB01 |
| Bupropion-naltrexone (Mysimba) | ATC: A08AA62 |
| Liraglutide (Saxenda) | ATC: A10BJ02 **AND** Item number*: 034982, 079356, 329833, 439932, 479008, 060677, 256177, 263604, 280312, 284144 |
| Semaglutide as WLD (Wegovy) | ATC: A10BJ06 **AND** Item number*: 191243, 386270, 409687, 418253, 534716 |
| Tirzepatide (Mounjaro) | ATC: A10BX16 |
| **COMORBIDITY GROUPS** | |
| **Cardiovascular Diseases** | |
| Hypertension | ICD-10: I10, I11, I12, I13, I15<br>ICPC2: K86, K87<br>ATC: C03A, C07, C08, C09 |
| Heart failure | ICD-10: I110, I130, I132, I50<br>ICPC2: K77 |
| Ischaemic heart disease | ICD-10: I20, I21, I23, I24, I25<br>ICPC2: K74, K75, K76 |
| Atrial fibrillation | ICD-10: I48<br>ICPC2: K78 |
| **Endocrine Disorders** | |
| Type 2 diabetes (T2D) | ICD-10: E11; ICPC2: T90 |
| Type 1 diabetes (T1D) | ICD-10: E10; ICPC2: T89 |
| Hyperlipidaemias | ICD-10: E78; ICPC2: T93; ATC: C10 |
| **Mental Disorders** | |
| ADHD | ICD-10: F90; ICPC2: P81 |
| Anxiety | ICD-10: F40, F41, F42: ICPC2: P74 |
| Depression | ICD-10: F32, F33, F34; ICPC2: P76 |
| Disorders due to psychoactive substance use (including alcohol) | ICD-10: F10, F11, F12, F13, F14, F15, F16, F18, F19, G312, G621, G721, I426, K292, K70, K860, O354, P043, Q860; ICPC2: P15, P16, P18, P19 |

| | |
|---|---|
| **Other Disorders** | |
| Sleep apnoea | ICD-10: G473 **AND** NCSP: EBGC02, ENC40, ENC45, GXAV37, GXAV38, GXAV40 |
| Stress incontinence | ICD-10: N393; ICPC2: U04 |
| Arthrosis | ICD-10: M15, M16, M17, M18, M19; ICPC2: L89, L90, L91 |
| Back pain | ICD-10: M54; ICPC2: L02, L84 |
| **Bariatric Surgery** | |
| Bariatric surgery | NCSP: JDF10, JDF11, JDF20, JDF21, JDF40, JDF41, JDC20, JDC21, JDD00, JDD01, JDE10, JDF50, JDF51, JDF96. JDF97, JDF98 |
| **COMEDICATION GROUPS** | |
| **Nervous System Medications** | |
| Antipsychotic | ATC: N05A |
| Antidepressants | ATC: N06A |
| Psychostimulants, agents used for ADHD and nootropics | ATC: N06BA |
| **Cardiovascular Medications** | |
| Cardiovascular | ATC: B01A, C01A, C01B, C01D, C01E, C03, C04, C07, C08, C09 |
| Lipid modifying agents | ATC: C10 |
| Antihypertensive | ATC: C03A, C07, C08, C09 |
| **Chronic Pain Medications** | |
| Anti-inflammatory and antirheumatic products | ATC: M01 |
| Opioids | ATC: N02A |
| Other analgesics and antipyretics | ATC: N02B |
| Paracetamol | ATC: N02BE |
| **Other Medications** | |
| Glucose lowering medications (except GLP-1 receptor agonists for obesity) | ATC: A10 |
| Medications for treatment of chronic lung diseases | ATC: R03A, R03B, R03D |
| **COMORBIDITY COUNT (10 DISTINCT GROUPS COUNTED)** | |
| **1) Cardiovascular diseases** | |

Hypertension, heart failure, ischaemic heart disease, atrial fibrillation combined (as defined above)

**2) Diabetes**

T1D and T2D diabetes combined (as defined above)

**3) Neoplasms**

Any ICD-10 code starting with “C”

**4) Sleep apnoea**

As defined above

**5) Liver disorders**

ICD-10: K72, K74, K758, K76, ICPC2 code D97

**6) Chronic obstructive pulmonary disease**

ICD-10: J41-J47; ICPC2: R95, R96; ATC: R03A, R03B, R03D

**7) Chronic kidney disease**

ICD-10; N18, N19

**8) Arthrosis/back pain**

Arthrosis and back pain combined, as defined above.

**9) Hypothyroidism/polycystic ovarian disease/ disorders of lipoprotein metabolism and other lipidaemias**

ICD-10: E03, E282; ICPC2 T86; ATC H03AA01

**10) Mental disorders**

Any mental disorder (as defined above)

#### Table S2. Comorbidity Proportions (%) among weight loss drug users stratified by sex and age groups (Sema: Semaglutide, Lira: Liraglutide, Tirz: Tirzepatide, B-N: Bupropion-naltrexone, Orli: Orlistat)

| | Women | | | | | | Men | | | | | |
|---|---|---|---|---|---|---|---|---|---|---|---|---|
| Age Group | Controls | Sema | Lira | Tirz | B-N | Orli | Controls | Sema | Lira | Tirz | B-N | Orli |
| Hypertension | | | | | | | | | | | | |
| 18-39 | 4.6 | 10 | 12 | 14 | 11 | 15 | 3.1 | 13 | 16 | 19 | 14 | 13 |
| 40-59 | 16 | 29 | 33 | 34 | 32 | 34 | 18 | 43 | 50 | 42 | 48 | 52 |
| 60-74 | 38 | 58 | 63 | 64 | 60 | 60 | 46 | 74 | 80 | 76 | 75 | 77 |
| Heart failure | | | | | | | | | | | | |
| 18-39 | 0.0 | 0.1 | 0.3 | - | 0.0 | - | 0.1 | 0.2 | 0.3 | - | 0.4 | - |
| 40-59 | 0.3 | 0.3 | 0.4 | 0.5 | 0.3 | - | 0.6 | 1.3 | 2.3 | 0.8 | 1.1 | 2.6 |
| 60-74 | 1.1 | 1.4 | 2.3 | 2.5 | 1.2 | 0.6 | 2.5 | 4.2 | 5.7 | 5.1 | 4.0 | 9.1 |
| Ischaemic heart disease | | | | | | | | | | | | |
| 18-39 | 0.0 | 0.1 | 0.1 | - | 0.1 | - | 0.1 | 0.4 | - | 0.4 | 0.7 | 1.0 |
| 40-59 | 0.7 | 1.2 | 2.3 | 1.9 | 1.3 | 2.5 | 2.5 | 4.7 | 6.5 | 5.2 | 4.7 | 6.5 |
| 60-74 | 3.2 | 4.8 | 6.0 | 6.2 | 5.2 | 6.1 | 9.7 | 14 | 19 | 15 | 14 | 16 |
| Atrial fibrillation | | | | | | | | | | | | |
| 18-39 | 0.1 | 0.1 | 0.2 | 0.2 | 0.1 | - | 0.1 | 0.4 | 0.8 | - | 0.4 | - |
| 40-59 | 0.4 | 0.7 | 0.7 | 0.7 | 0.6 | 0.8 | 1.0 | 2.6 | 3.1 | 2.1 | 2.1 | 1.7 |
| 60-74 | 2.2 | 3.8 | 6.2 | 5.9 | 3.4 | 2.2 | 5.3 | 10 | 11 | 9.1 | 7.6 | 13 |
| T1D | | | | | | | | | | | | |
| 18-39 | 0.6 | 0.6 | 1.3 | 0.2 | 1.4 | 2.3 | 0.8 | 0.7 | 2.2 | - | 1.4 | 3.0 |
| 40-59 | 0.9 | 0.5 | 1.5 | 0.8 | 1.1 | 0.4 | 1.2 | 0.8 | 2.6 | 0.9 | 1.9 | 2.6 |
| 60-74 | 1.1 | 0.6 | 1.7 | 1.0 | 1.4 | 2.2 | 1.6 | 1.0 | 2.4 | 1.2 | 1.9 | 1.3 |
| T2D | | | | | | | | | | | | |
| 18-39 | 0.7 | 1.2 | 2.1 | 2.2 | 2.1 | 3.3 | 0.8 | 1.6 | 3.1 | 2.6 | 2.6 | 2.0 |
| 40-59 | 3.8 | 2.9 | 4.5 | 5.5 | 5.6 | 5.8 | 5.5 | 5.0 | 8.5 | 7.6 | 9.0 | 14 |
| 60-74 | 8.7 | 7.4 | 9.5 | 9.6 | 11 | 8.8 | 14 | 12 | 14 | 17 | 15 | 22 |
| Hyperlipidaemias | | | | | | | | | | | | |
| 18-39 | 1.4 | 3.5 | 4.2 | 4.2 | 4.6 | 6.5 | 2.3 | 8.3 | 8.4 | 9.5 | 8.4 | 9.0 |
| 40-59 | 9.3 | 15 | 17 | 19 | 17 | 20 | 15 | 28 | 32 | 29 | 29 | 33 |
| 60-74 | 31 | 41 | 48 | 47 | 43 | 42 | 39 | 55 | 58 | 55 | 54 | 53 |
| ADHD | | | | | | | | | | | | |
| 18-39 | 4.3 | 7.0 | 8.1 | 7.6 | 7.2 | 7.2 | 4.0 | 7.5 | 12 | 8.8 | 9.1 | 10 |
| 40-59 | 1.7 | 2.6 | 3.1 | 3.0 | 2.5 | 3.5 | 1.6 | 2.5 | 3.0 | 3.1 | 2.6 | 3.9 |
| 60-74 | 0.3 | 0.6 | 0.8 | 1.7 | 0.5 | - | 0.4 | 0.6 | 0.4 | 1.6 | 0.5 | 1.3 |
| Anxiety | | | | | | | | | | | | |
| 18-39 | 6.8 | 11 | 14 | 16 | 12 | 15 | 3.4 | 6.2 | 8.4 | 6.2 | 7.4 | 6.0 |
| 40-59 | 4.1 | 5.9 | 7.9 | 8.2 | 6.3 | 8.7 | 2.6 | 3.5 | 4.8 | 3.9 | 3.8 | 3.5 |
| 60-74 | 3.3 | 4.0 | 6.6 | 5.2 | 3.6 | 5.0 | 1.8 | 2.3 | 4.5 | 3.6 | 2.1 | 5.2 |
| Depression | | | | | | | | | | | | |
| 18-39 | 9.9 | 19 | 28 | 27 | 20 | 22 | 6.2 | 12 | 19 | 13 | 14 | 13 |
| 40-59 | 8.2 | 13 | 19 | 19 | 14 | 17 | 5.2 | 8.3 | 12 | 12 | 10 | 11 |
| 60-74 | 5.7 | 8.5 | 11 | 14 | 8.8 | 10 | 3.5 | 5.2 | 11 | 5.1 | 5.9 | 10 |
| Drug-related disorders | | | | | | | | | | | | |
| 18-39 | 1.5 | 2.3 | 2.7 | 1.8 | 2.3 | 4.6 | 3.1 | 3.6 | 5.6 | 2.9 | 3.1 | 3.0 |
| 40-59 | 1.4 | 1.7 | 3.2 | 2.5 | 1.5 | 2.9 | 2.9 | 2.7 | 3.8 | 3.5 | 2.6 | 7.8 |
| 60-74 | 1.5 | 1.6 | 2.9 | 2.7 | 1.3 | 2.8 | 2.7 | 2.7 | 6.1 | 3.6 | 3.4 | 3.9 |
| Sleep apnoea | | | | | | | | | | | | |
| 18-39 | 0.3 | 1.8 | 3.4 | 2.5 | 2.7 | 3.3 | 1.3 | 8.1 | 12 | 11 | 10 | 7.0 |
| 40-59 | 1.5 | 4.8 | 6.8 | 6.7 | 5.6 | 5.2 | 3.6 | 14 | 23 | 17 | 18 | 15 |
| 60-74 | 2.4 | 8.8 | 13 | 15 | 11 | 7.2 | 4.9 | 18 | 26 | 22 | 21 | 25 |

| | | | | | | | | | | | | |
|---|---|---|---|---|---|---|---|---|---|---|---|---|
| Stress incontinence | | | | | | | | | | | | |
| 18-39 | 0.9 | 2.1 | 2.0 | 3.6 | 2.5 | 2.9 | 0.1 | 0.2 | - | - | 0.4 | - |
| 40-59 | 2.7 | 4.7 | 6.1 | 6.7 | 5.5 | 7.9 | 0.3 | 0.4 | 0.1 | 0.5 | 0.4 | 0.4 |
| 60-74 | 3.0 | 5.9 | 7.1 | 7.1 | 7.6 | 4.4 | 1.1 | 1.8 | 3.7 | 2.0 | 1.8 | 1.3 |
| Arthrosis | | | | | | | | | | | | |
| 18-39 | 0.5 | 1.2 | 1.3 | 0.5 | 1.5 | 1.0 | 0.6 | 1.2 | 0.6 | 1.8 | 1.5 | 1.0 |
| 40-59 | 5.1 | 10 | 13 | 12 | 13 | 11 | 3.6 | 7.6 | 12 | 7.6 | 10 | 13 |
| 60-74 | 14 | 26 | 32 | 30 | 28 | 24 | 8.8 | 19 | 22 | 19 | 20 | 17 |
| Back pain | | | | | | | | | | | | |
| 18-39 | 8.0 | 15 | 19 | 24 | 17 | 18 | 7.5 | 14 | 17 | 20 | 17 | 23 |
| 40-59 | 10 | 15 | 20 | 21 | 19 | 23 | 9.6 | 14 | 20 | 18 | 17 | 18 |
| 60-74 | 10 | 16 | 17 | 19 | 18 | 20 | 9.3 | 15 | 15 | 17 | 17 | 12 |

Controls: Shown for Semaglutide only.

#### Table S3. Comedication Proportions (%) among weight loss drug users stratified by sex and age groups (Sema: Semaglutide, Lira: Liraglutide, Tirz: Tirzepatide, B-N: Bupropion-naltrexone, Orli: Orlistat)

| | Women | | | | | | Men | | | | | |
|---|---|---|---|---|---|---|---|---|---|---|---|---|
| Age Group | Controls | Sema | Lira | Tirz | B-N | Orli | Controls | Sema | Lira | Tirz | B-N | Orli |
| Antipsychotic | | | | | | | | | | | | |
| 18-39 | 3.0 | 6.6 | 8.6 | 11 | 5.8 | 11 | 2.7 | 5.8 | 6.5 | 8.4 | 5.0 | 6.0 |
| 40-59 | 3.8 | 5.8 | 9.3 | 9.9 | 5.8 | 11 | 3.3 | 4.7 | 6.5 | 6.3 | 4.9 | 7.4 |
| 60-74 | 4.0 | 5.9 | 7.7 | 13 | 5.2 | 7.2 | 3.0 | 3.8 | 4.9 | 4.7 | 4.7 | 5.2 |
| Antidepressants | | | | | | | | | | | | |
| 18-39 | 8.6 | 20 | 28 | 30 | 21 | 26 | 4.6 | 12 | 16 | 15 | 13 | 15 |
| 40-59 | 12 | 21 | 28 | 29 | 24 | 28 | 6.2 | 12 | 14 | 17 | 14 | 16 |
| 60-74 | 13 | 21 | 27 | 27 | 22 | 29 | 6.6 | 11 | 15 | 15 | 12 | 16 |
| Psychostimulants | | | | | | | | | | | | |
| 18-39 | 3.0 | 4.5 | 5.1 | 4.9 | 4.3 | 4.9 | 2.5 | 4.9 | 7.0 | 7.3 | 4.9 | 3.0 |
| 40-59 | 1.3 | 2.0 | 2.6 | 2.6 | 1.9 | 2.5 | 1.2 | 2.0 | 2.6 | 2.4 | 1.9 | 2.6 |
| 60-74 | 0.3 | 0.5 | 0.4 | 1.0 | 0.4 | - | 0.3 | 0.6 | 0.4 | 2.4 | 0.5 | 1.3 |
| Cardiovascular | | | | | | | | | | | | |
| 18-39 | 5.2 | 12 | 14 | 16 | 13 | 16 | 2.9 | 11 | 15 | 18 | 13 | 13 |
| 40-59 | 17 | 31 | 36 | 37 | 34 | 36 | 19 | 43 | 50 | 44 | 48 | 51 |
| 60-74 | 40 | 62 | 69 | 68 | 65 | 65 | 51 | 78 | 85 | 78 | 78 | 74 |
| Lipid modifying agents | | | | | | | | | | | | |
| 18-39 | 0.7 | 1.8 | 2.3 | 3.1 | 2.7 | 3.6 | 1.3 | 4.8 | 6.5 | 5.9 | 5.3 | 5.0 |
| 40-59 | 7.4 | 12 | 14 | 16 | 14 | 17 | 13 | 24 | 28 | 24 | 26 | 27 |
| 60-74 | 27 | 38 | 44 | 43 | 39 | 39 | 36 | 52 | 56 | 53 | 50 | 51 |
| Antihypertensives | | | | | | | | | | | | |
| 18-39 | 3.3 | 7.7 | 10 | 11 | 8.8 | 12 | 2.3 | 9.8 | 12 | 16 | 12 | 11 |
| 40-59 | 14 | 25 | 30 | 30 | 28 | 28 | 16 | 39 | 46 | 38 | 44 | 46 |
| 60-74 | 34 | 54 | 58 | 59 | 57 | 58 | 43 | 71 | 79 | 71 | 72 | 70 |
| Anti-inflammatory drugs | | | | | | | | | | | | |
| 18-39 | 17 | 33 | 41 | 46 | 38 | 39 | 12 | 27 | 33 | 37 | 30 | 34 |
| 40-59 | 27 | 44 | 54 | 58 | 52 | 55 | 20 | 36 | 40 | 45 | 41 | 46 |
| 60-74 | 26 | 43 | 47 | 53 | 49 | 45 | 20 | 34 | 36 | 47 | 38 | 39 |
| Opioids | | | | | | | | | | | | |
| 18-39 | 9.4 | 21 | 29 | 29 | 23 | 26 | 7.6 | 18 | 22 | 27 | 19 | 24 |
| 40-59 | 14 | 26 | 35 | 39 | 29 | 34 | 12 | 23 | 30 | 32 | 25 | 30 |
| 60-74 | 17 | 29 | 36 | 38 | 29 | 34 | 15 | 27 | 34 | 39 | 28 | 23 |
| Paracetamol | | | | | | | | | | | | |
| 18-39 | 10 | 24 | 30 | 31 | 28 | 24 | 6.4 | 16 | 23 | 21 | 19 | 28 |
| 40-59 | 21 | 37 | 46 | 45 | 46 | 46 | 13 | 25 | 31 | 31 | 32 | 34 |
| 60-74 | 27 | 48 | 53 | 51 | 53 | 55 | 19 | 34 | 45 | 46 | 40 | 34 |
| Other analgesics | | | | | | | | | | | | |
| 18-39 | 11 | 25 | 31 | 33 | 29 | 26 | 6.8 | 17 | 25 | 22 | 20 | 29 |
| 40-59 | 22 | 39 | 47 | 47 | 47 | 48 | 13 | 26 | 33 | 34 | 34 | 37 |
| 60-74 | 28 | 49 | 54 | 52 | 54 | 56 | 20 | 36 | 45 | 47 | 42 | 40 |
| Glucose lowering drugs* | | | | | | | | | | | | |
| 18-39 | 1.8 | 6.1 | 8.4 | 9.9 | 7.1 | 9.8 | 1.2 | 2.0 | 4.8 | 2.9 | 3.1 | 3.0 |
| 40-59 | 3.9 | 3.5 | 5.2 | 7.9 | 5.5 | 6.8 | 5.4 | 4.4 | 7.6 | 7.9 | 7.8 | 12 |
| 60-74 | 7.8 | 5.3 | 6.8 | 9.9 | 8.7 | 8.3 | 13 | 9.7 | 13 | 16 | 12 | 16 |
| Chronic lung disease drugs | | | | | | | | | | | | |
| 18-39 | 6.5 | 13 | 15 | 21 | 14 | 16 | 5.0 | 11 | 11 | 14 | 13 | 17 |
| 40-59 | 10 | 17 | 22 | 23 | 20 | 22 | 7.4 | 14 | 17 | 16 | 15 | 18 |
| 60-74 | 15 | 26 | 29 | 27 | 28 | 25 | 12 | 21 | 26 | 26 | 24 | 23 |

Controls: Shown for Semaglutide only.

### Reimbursement regulations in Norway

Norway has a tax-financed universal healthcare system with contacts at public hospitals free of charge for patients. Further, there is low copayment for visits to general practitioners in primary care and for drugs that are reimbursable for treatment of chronic conditions, with a total yearly patient compayment around 270€.

#### Orlistat

Orlistat is a peripherally acting antiobesity medication. Marketed in 2000 in Norway. Orlistat is reimbursed to patients with BMI ≥40 or BMI ≥35 with one additional disease.

Monthly cost is around 900NOK (~75€ a month).

| **Obesity related diagnosis:** | **ICD-10** | **ICPC-2** |
|---|---|---|
| Candidiasis of skin and nail | B37.2 | S75 |
| Hypothyroidism, unspecified | E03.9 | T86 |
| Polycystic ovarian syndrome (PCOS) | E28.2 | T99 |
| Disorders of lipoprotein metabolism and other lipidaemia | E78 | T93 |
| Other specified metabolic disorders | E88.8 | T99 |
| Hyperuricemia without signs of inflammatory arthritis and tophaceous disease | E79.0 | T99 |
| Type 2 diabetes | E11 | T90 |
| Mood (affective) disorders | F30, F31, F32, F33, F34, F38 | P76 |
| Other anxiety disorders | F41 | P74 |
| Overeating associated with other psychological disturbances | F50.4 | P86 |
| Hypertension | I10, I11, I12, I13, I15 | K86, K87 |
| Acute myocardial infarction | I21 | K75 |
| Subsequent myocardial infarction | I22 | K74, K76 |
| Chronic ischemic heart disease | I25 | K76 |
| Other pulmonary heart diseases | I27 | K82 |
| Heart failure | I50 | K77 |
| Stroke, not specified as haemorrhage or infarction | I64 | K90 |
| Transient cerebral ischemic attack, unspecified | G45.9 | K89 |
| Obstructive sleep apnoea (OSA) | G47.3 | P06 |
| Gastro-oesophageal reflux disease (GERD) | K21 | D84 |
| Hepatic failure, not elsewhere classified | K72 | D97 |
| Fibrosis and cirrhosis of the liver | K74 | D97 |
| Other diseases of liver | K76 | D97 |
| Idiopathic gout | M10.0 | T92 |
| Polyosteoarthritis | M15 | L91 |
| Osteoarthritis of hip (Coxarthrosis) | M16 | L89 |

| | | |
|---|---|---|
| Osteoarthritis of the knee (Gonarthrosis) | M17 | L90 |
| Chronic kidney disease | N18 | U99 |
| Stress incontinence | N39.3 | U04 |

https://www.helsedirektoratet.no/rundskriv/kapittel-5-stonad-ved-helsetjenester/vedlegg-1-til--5-14-legemiddellisten/virkestoffer/orlistat

#### Bupropion-naltrexone

Bupropion-naltrexone is preapproved as reimbursable by the Norwegian health authorities. From 2017 to 2020 it was only reimbursed for individuals with BMI a ≥40 kg/m2 or BMI ≥35 kg/$m^2$ with comorbidities, such as hypertension, type 2 diabetes, and obstructive sleep apnoea. In 2020, the reimbursement criteria changed, and the list of eligible comorbid conditions increased to 29 conditions (see table with diagnosis codes) for those with a BMI between 35 and 40 kg/$m^2$. Any physician can prescribe it to adults. The monthly cost is around 1300NOK (~110€).

| Obesity related diagnosis: | ICD-10 | ICPC-2 |
|---|---|---|
| Candidiasis of skin and nail | B37.2 | S75 |
| Hypothyroidism, unspecified | E03.9 | T86 |
| Polycystic ovarian syndrome (PCOS) | E28.2 | T99 |
| Disorders of lipoprotein metabolism and other lipidaemia | E78 | T93 |
| Other specified metabolic disorders | E88.8 | T99 |
| Hyperuricemia without signs of inflammatory arthritis and tophaceous disease | E79.0 | T99 |
| Type 2 diabetes | E11 | T90 |
| Mood (affective) disorders | F30, F31, F32, F33, F34, F38 | P76 |
| Other anxiety disorders | F41 | P74 |
| Overeating associated with other psychological disturbances | F50.4 | P86 |
| Hypertension | I10, I11, I12, I13, I15 | K86, K87 |
| Acute myocardial infarction | I21 | K75 |
| Subsequent myocardial infarction | I22 | K74, K76 |
| Chronic ischemic heart disease | I25 | K76 |
| Other pulmonary heart diseases | I27 | K82 |
| Heart failure | I50 | K77 |
| Stroke, not specified as haemorrhage or infarction | I64 | K90 |
| Transient cerebral ischemic attack, unspecified | G45.9 | K89 |
| Obstructive sleep apnoea (OSA) | G47.3 | P06 |
| Gastro-oesophageal reflux disease (GERD) | K21 | D84 |
| Hepatic failure, not elsewhere classified | K72 | D97 |
| Fibrosis and cirrhosis of the liver | K74 | D97 |
| Other diseases of liver | K76 | D97 |
| Idiopathic gout | M10.0 | T92 |
| Polyosteoarthritis | M15 | L91 |

| | | |
|---|---|---|
| Osteoarthritis of hip (Coxarthrosis) | M16 | L89 |
| Osteoarthritis of the knee (Gonarthrosis) | M17 | L90 |
| Chronic kidney disease | N18 | U99 |
| Stress incontinence | N39.3 | U04 |

https://www.helsedirektoratet.no/rundskriv/kapittel-5-stonad-ved-helsetjenester/vedlegg-1-til--5-14-legemiddellisten/virkestoffer/naltrekson-og-bupropion

#### Liraglutide (Saxenda)

Liraglutide with indication for obesity (Saxenda) was introduced in the Norwegian market in 2019 and was reimbursed from 2020 to January 2023 for patients with BMI ≥35 and comorbidities. Individual applications for reimbursement were granted until 1st February 2023. The monthly cost is around 1 300NOK (110€).

| **Obesity related diagnosis:** | **ICD-10** | **ICPC-2** |
|---|---|---|
| Disorders of lipoprotein metabolism and other lipidaemia | E78 | T93 |
| Type 2 diabetes | E11 | T90 |
| Obstructive sleep apnoea (OSA) | G47.3 | P06 |
| Essential hypertension | I10 | K86 |
| Hypertensive heart disease | I11 | K86 |
| Hypertensive chronic kidney disease | I12 | K86 |
| Hypertensive heart and chronic kidney disease | I13 | K86 |
| Secondary hypertension | I15 | K87 |
| Chronic ischemic heart disease | I25 | K76 |
| Heart failure | I50 | K77 |
| Atherosclerosis | I70 | K92 |

#### Semaglutide (Wegovy)

Semaglutide Wegovy has been available in Norway from January 2023 but has never been reimbursed.

Only adult patients with a BMI over 50 who also have serious obesity related comorbidities (see list with diagnosis codes under) can be assessed for reimbursement, the applications must come from doctors working at the hospital. There special reimbursement rules for children 12 to 18 years old.

Prescribing Ozempic (semaglutide with type 2 diabetes indication) for obesity in Norway is outside the approved indication, prescriber takes on a special responsibility when it comes to assessing the benefit-risk ratio and follow-up with the patient regarding effects and side effects.

The cost for Wegovy in Norway is around 3 200NOK (~270 euro) per month in a dose of 2.4mg/weekly.

| **Obesity related diagnosis:** | **ICD-10** | **ICPC-2** |
|---|---|---|
| Disorders of lipoprotein metabolism and other lipidaemia | E78 | T93 |
| Type 2 diabetes | E11 | T90 |
| Obstructive sleep apnoea (OSA) | G47.3 | P06 |
| Essential hypertension | I10 | K86 |
| Hypertensive heart disease | I11 | K86 |
| Hypertensive chronic kidney disease | I12 | K86 |
| Hypertensive heart and chronic kidney disease | I13 | K86 |
| Secondary hypertension | I15 | K87 |
| Chronic ischemic heart disease | I25 | K76 |
| Heart failure | I50 | K77 |
| Atherosclerosis | I70 | K92 |

https://www.helsedirektoratet.no/rundskriv/kapittel-5-stonad-ved-helsetjenester/vedlegg-1-til--5-14-legemiddellisten/virkestoffer/semaglutid

#### Tirzepatide (Mounjaro)

Tirzepatide (Mounjaro) has been available from November 2024 in Norway and is not reimbursed.

The cost for Mounjaro in Norway is around 4 700NOK (400 euros) per month in a dose of 7,5 to 10 mg/weekly and 4 900NOK (415 euros) per month in a weekly dose of 12,5 to 15 mg.
